# Evaluation of Sex-Related Differences in Cerebrovascular Bypass Patency: An Institutional Review of 357 Direct Cerebral Bypasses

**DOI:** 10.1101/2024.09.22.24314174

**Authors:** Laura Stone McGuire, Tatiana Abou-Mrad, Xinjian Du, Ali Alaraj, Sepideh Amin-Hanjani, Gursant Atwal, Fady T. Charbel

## Abstract

**Introduction:** Demographics and comorbid conditions play a role in vascular health, yet their specific impact on cerebrovascular bypass patency remains unclear.

**Methods:** An institutional database of 357 patients with intracranial bypass procedures between 08/2001-05/2022 was retrospectively reviewed. Patients with bypass for all causes (e.g., aneurysm, atherosclerotic disease, Moyamoya disease) were included. Medical history, surgical technique, and flow-related measurements (intraoperatively and on quantitative MRA at follow-up) were compared across biological sex and in relation to bypass patency.

**Results:** Of 357 patients, 141 were male (39.5%) and 216 were female (60.5%) with average age 49.0+/-16.7. Bypass patency at last follow-up was 84.4% for men vs. 69.2% for women (p=0.001). Significant differences were seen in underlying diagnoses, with more aneurysm and Moyamoya cases represented in female sex (p<0.001); irrespective of diagnosis, lower patency rates were seen in women when considering bypass for aneurysm (p=0.032), Moyamoya disease (p=0.035), and for atherosclerotic disease (p=0.159). Medical comorbidities were seen at higher rates in men, with comorbidity score 2.7 vs. 2.1 (p<0.001). Cut flow was higher in men 59.2 vs. 51.1 (p=0.028), but no significant differences were seen in intraoperative bypass flow, cut flow index (CFI), or follow-up QMRA. After removing cases using interposition grafts, similar differences were redemonstrated. Propensity score matched analysis found females have a 2.71 higher chance of bypass occlusion after adjusting for CFI (p=0.017, 95% CI: 1.19-6.18).

**Conclusion:** Biological sex appears to play a significant role in bypass patency, across diagnoses. Women were significantly less likely to have patent bypasses at last follow-up, despite having less medical comorbidities than men and despite having similar intraoperative and perioperative flows. Further study is required to better elucidate the influence of sex on long-term bypass patency.

## INTRODUCTION

The role of sex differences in cerebrovascular diseases has gained increasing attention in recent years. Growing evidence indicates that the etiology, clinical course, and outcomes of cerebrovascular conditions, such as stroke, may differ significantly between men and women.^1–3^ For instance, women exhibit a higher incidence of intracranial aneurysms (IAs), along with a markedly increased incidence of subarachnoid hemorrhage compared to men, who, in contrast, demonstrate higher rates of hemorrhagic stroke and large artery atherosclerosis.^4–7^ These differences have profound implications for both primary and secondary prevention strategies, as well as treatment modalities.

Some reproductive factors, such as pregnancy and menopause, predispose women to an increased risk of cerebrovascular events.^3,8^ However, men more frequently present with traditional vascular risk factors (such as hypertension, dyslipidemia, and smoking), which are strongly associated with a higher incidence of severe stroke, among other cerebrovascular pathologies.^9–11^ This underscores the need for sex-specific research to elucidate the underlying pathophysiological mechanisms. As the impact of sex on cerebrovascular health extends beyond epidemiological differences, it is crucial to consider how these factors influence clinical outcomes, as tailored clinical care could significantly enhance the outcomes of cerebrovascular interventions.

In the context of intracranial bypass surgery, sex-specific differences remain underexplored, particularly regarding long-term bypass patency. This procedure is a critical intervention for several cerebrovascular diseases, including IAs, Moyamoya disease (MMD), and atherosclerotic conditions, where restoring and augmenting cerebral blood flow is paramount. Understanding the influence of sex on the success of this intervention is essential for refining surgical techniques, optimizing peri and postoperative management, and improving patient outcomes. This study aims to provide an exploratory assessment of the impact of sex on intracranial bypass outcomes.

## METHODS

### Patient Selection

An institutional database of 357 patients with intracranial bypass procedures between August 2001 and May 2022 was retrospectively reviewed following institutional review board approval. The requirement for patient consent was waived due to the retrospective nature of the study. Patients with direct bypass for all causes (e.g., aneurysm, atherosclerotic disease, MMD) were included. Medical history, surgical technique, flow-related measurements (intraoperatively and on quantitative MRA at follow-up), and follow-up data were compared across biological sex and in relation to bypass patency. Medical history was collected from the electronic medical record, including the following comorbidities (hypertension, hyperlipidemia, diabetes mellitus, coronary artery disease, peripheral vascular disease, chronic kidney disease, prior stroke/TIA, and tobacco use), and the comorbidity score was a sum of all present comorbid conditions in the patient at the time of surgery. The decision to perform a bypass was at the discretion of the treating surgeons. Standard indications for bypass at this institution include symptomatic MMD with hemodynamic compromise, failure of maximal medial therapy with hemodynamic compromise for ICAD, and aneurysms not amenable to direct clipping or endovascular strategies. Bypasses were performed under general anesthesia using previously described techniques.^12^

### Blood Flow Measurements

Flow measurements were performed intraoperatively using the intracranial micro-flow probe via transit-time ultrasound technology (Charbel Micro-Flowprobe; Transonics Systems Inc., Ithaca, NY, USA).^13–14^ Cut flow index (CFI, ratio of intraoperative bypass flow to initial flow of the donor vessel), which is a demonstrated predictor of bypass patency in occlusive disease was noted.^15–16^ Postoperatively, bypass patency was assessed by digital subtraction angiography and by follow-up digital subtraction angiography, computed tomography angiography, or magnetic resonance angiography.

Blood flow within the bypass was measured with QMRA via commercially available NOVA software (Non-invasive Optimal Vessel Analysis, VasSol, Inc., River Forest, Illinois), which has been described and validated in several cerebrovascular pathologies.^17–25^ Postoperative bypass flow data via QMRA was available in 165 patients within 30 days of surgery.

Once blood flow and vessel diameter were measured, wall shear stress (WSS) was calculated using the Hagen-Poiseuille equation:

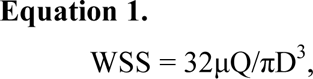

where WSS is in dynes/cm^2^, Q is the volumetric flow rate in mL/sec, and D is the vessel diameter in cm; μ is the blood viscosity in poise and was assumed to be constant (0.035 poise). This method was previously described by Zhao et al.^25^

In addition, using QMRA data, pulsatility index (PI) and resistivity index (RI) were calculated as follows:

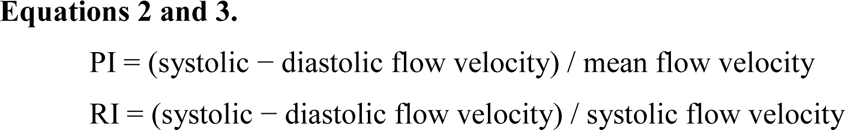

### Statistical Analysis

All statistical analyses were performed using SPSS version 29 (IBM, Inc., Armonk, New York), using 2-sided tests with significance level α=0.05. Demographic, clinical, procedural, and hemodynamic characteristics were compared between male and female patients. Given the exploratory nature of the analysis, adjustment for multiple comparisons were not applied. Student t-test with Levene’s test for equality of variances was used to compare means of continuous data between 2 groups. Chi-square test was used to compare proportions among categorical data in 2 groups, and the Fisher’s exact test was used if sample size was small. 2-way ANOVA was conducted to assess multiple hemodynamic parameters to compare patients with and without patent bypasses on last follow-up, per patient sex. Propensity score matched analysis was performed to compare females and males (n=114, n=114), matching comorbidities and risk factors (hypertension, hyperlipidemia, diabetes, coronary artery disease, prior stroke or TIA, tobacco use, and race/ethnicity), to assess bypass patency (**Supplemental Table 1**). Only patients with complete data across listed variables were included in propensity scoring.

## RESULTS

### Initial Analysis of Bypass Patency

Of 357 patients, 141 were male (39.5%) and 216 were female (60.5%) with average age 49.0+/-16.7. 349 patients had follow-up data. **Table 1** summarizes the initial analysis comparing patients with occluded vs. patent bypasses with regards to demographic data, comorbidities, primary diagnosis, bypass descriptions, and hemodynamic parameters. Comparing patent and occluded bypasses, of all types and indications, women had significantly higher rates of occlusion (30.8% vs. 15.6%, p=0.001). All other characteristics, including cardiovascular risk factors, were shown at similar rates in both patent and occluded bypasses. Regarding flow-related and hemodynamic parameters, only CFI differed significantly between patent and occluded bypasses (0.92 vs 0.69, p=0.003). Patent bypasses showed a trend for higher QMRA flow measurement, particularly on last follow-up imaging, and for larger diameters, albeit non-significant.

**Table 1.** Characteristics of patients and their risk factors, comparing patent and occluded bypasses (at last follow-up). mRS = modified Rankin scale; BMI = body mass index; HTN = hypertension; DM = diabetes mellitus; CAD = coronary artery disease; PAD = peripheral arterial disease; CKD = chronic kidney disease; TIA = transient ischemic attack; STA = superficial temporal artery; MCA = middle cerebral artery; NOVA = non-invasive optimal vessel analysis.

|  | Not patent (n=87) | Patent (n=262) | p-value |
| --- | --- | --- | --- |
| Age | 49.43 +/- 16.06 | 48.70 +/- 17.11 | 0.730 |
| Sex |  |  | **0.001** |
| Male | 21 / 87 (15.6%) | 114 / 262 (84.4%) |  |
| Female | 66 / 87 (30.8%) | 148 / 262 (69.2%) |  |
| Diagnosis |  |  | 0.819 |
| Aneurysm | 27 / 87 (23.7%) | 87 / 262 (76.3%) |  |
| Moyamoya | 32 / 87 (26.2%) | 90 / 262 (73.8%) |  |
| Atherosclerotic | 27 / 87 (25.2%) | 80 / 262 (74.8%) |  |
| Other | 1 / 87 (25.0%) | 3 / 262 (75.0%) |  |
| mRS at surgery |  |  | 0.182 |
| 0 | 1 / 87 (1.1%) | 10 / 257 (3.9%) |  |
| 1 | 27 / 87 (31.0%) | 65 / 257 (25.3%) |  |
| 2 | 42 / 87 (48.3%) | 105 / 257 (40.9%) |  |
| 3 | 15 / 87 (17.2%) | 55 / 257 (21.4%) |  |
| 4 | 1 / 87 (1.1%) | 16 / 257 (6.2%) |  |
| 5 | 1 / 87 (1.1%) | 6 / 257 (2.3%) |  |
| Race/Ethnicity |  |  | 0.427 |
| White | 49 / 82 (59.8%) | 128 / 247 (51.8%) |  |
| AfricanAmerican | 16 / 82 (19.5%) | 41 / 247 (16.6%) |  |
| Hispanic | 4 / 82 (4.9%) | 23 / 247 (9.3%) |  |
| Asian | 4 / 82 (4.9%) | 14 / 247 (5.7%) |  |
| Other | 9 / 82 (11.0%) | 41 / 247 (16.6%) |  |
| Weight (kg) | 85.08 +/- 23.89 | 85.44 +/- 23.27 | 0.908 |
| BMI | 30.24 +/- 8.08 | 29.02 +/- 8.12 | 0.275 |
| Comorbidities |  |  |  |
| HTN | 52 / 82 (63.4%) | 158 / 249 (63.5%) | 0.995 |
| HLD | 31 / 82 (37.8%) | 98 / 249 (39.4%) | 0.803 |
| DM | 19 / 82 (23.2%) | 59 / 249 (23.7%) | 0.923 |
| CAD | 12 / 82 (14.6%) | 34 / 249 (13.7%) | 0.824 |
| PAD | 3 / 82 (3.7%) | 12 / 249 (4.8%) | 1.000 |
| CKD | 3 / 82 (3.7%) | 5 / 248 (2.0%) | 0.416 |
| Stroke/TIA | 37 / 82 (45.1%) | 112 / 249 (45.0%) | 0.982 |
| Tobacco | 37 / 82 (45.1%) | 106 / 249 (42.6%) | 0.686 |
| Comorbidity score | 2.37 +/- 1.45 | 2.35 +/- 1.58 | 0.956 |
| Medications |  |  |  |
| Plavix | 13 / 82 (15.9%) | 35 / 249 (14.1%) | 0.688 |
| Aspirin | 79 / 82 (96.3%) | 235 / 249 (94.4%) | 0.485 |
| Statin | 35 / 82 (42.7%) | 104 / 249 (41.8%) | 0.884 |
| Anastomosis sites | 1.30 +/- 0.51 | 1.30 +/- 0.46 | 0.948 |
| Bypass type |  |  | 0.944 |
| STA-MCA | 62 / 87 (71.3%) | 191 / 262 (72.9%) |  |
| Graft-MCA | 11 / 87 (12.6%) | 27 / 262 (10.3%) |  |
| Other-anterior | 5 / 87 (5.7%) | 15 / 262 (5.7%) |  |
| Other-posterior | 9 / 87 (10.3%) | 29 / 262 (11.1%) |  |
| **Intraoperative Measurements** |  |  |  |
| Cut flow | 54.18 +/- 28.64 | 54.45 +/- 29.31 | 0.951 |
| Bypass flow | 46.90 +/- 43.19 | 50.48 +/- 35.02 | 0.481 |
| Cut flow index | 0.69 +/- 0.43 | 0.92 +/- 0.51 | **0.003** |

| QMRA Measurements |  |  |  |
| --- | --- | --- | --- |
| NOVA first flow | 65.08 +/- 34.10 | 70.41 +/- 40.28 | 0.537 |
| NOVA last flow | 28.36 +/- 22.07 | 49.15 +/- 34.46 | 0.068 |
| Pulsatility index | 0.33 +/- 0.26 | 0.32 +/- 0.16 | 0.829 |
| Resistivity index | 0.27 +/- 0.16 | 0.25 +/- 0.15 | 0.617 |
| Wall shear stress | 1.53 +/- 1.23 | 1.33 +/- 0.78 | 0.258 |
| Bypass diameter | 2.64 +/- 0.54 | 2.87 +/- 0.62 | 0.056 |

### Sex-Related Differences in Bypass Characteristics and Patency

Bypass patency at last follow-up was 84.4% for men vs. 69.2% for women (p=0.001), with a similar length of follow-up was (1.81 vs. 2.28 years, p=0.138). Significant differences were seen in underlying diagnoses, with more aneurysm and Moyamoya cases represented in female sex (p<0.001). When looking at these specific indications for bypass, lower patency rates were seen in women when considering bypass for aneurysm (p=0.032), MMD (p=0.035), and for atherosclerotic disease (p=0.159). Bypass type did significantly vary, and the difference appeared to be driven by higher interposition graft usage in women (p=0.005). **Table 2** summarizes these findings. Repeat analyses were performed to exclude interposition grafts and study STA-MCA bypasses only, the findings of which are summarized in **Table 3** with similar reported differences between men and women.

**Table 2.** Characteristics of patients with all bypass types, comparing male and female. mRS = modified Rankin scale; BMI = body mass index; HTN = hypertension; DM = diabetes mellitus; CAD = coronary artery disease; PAD = peripheral arterial disease; CKD = chronic kidney disease; TIA = transient ischemic attack; STA = superficial temporal artery; MCA = middle cerebral artery; NOVA = non-invasive optimal vessel analysis.

|  | Male (n=141) | Female (n=216) | p-value |
| --- | --- | --- | --- |
| Age | 49.81 +/- 17.51 | 48.41 +/- 16.13 | 0.443 |
| Length of follow-up (year) | 1.81 +/- 2.51 | 2.28 +/- 3.09 | 0.138 |
| Bypass patent (at last follow-up) | 114 / 135 (84.4%) | 148 / 214 (69.2%) | **0.001** |
| Diagnosis |  |  | **<0.001** |
| Aneurysm | 37 / 141 (26.2%) | 79 / 215 (36.7%) |  |
| Moyamoya | 38 / 141 (27.0%) | 85 / 215 (39.5%) |  |
| Atherosclerotic | 61 / 141 (43.3%) | 47 / 215 (21.9%) |  |
| Other | 5 / 141 (3.5%) | 4 / 215 (1.8%) |  |
| mRS at surgery |  |  | 0.413 |
| 0 | 5 / 139 (3.6%) | 7 / 212 (3.3%) |  |
| 1 | 36 / 139 (25.9%) | 59 / 212 (27.8%) |  |
| 2 | 54 / 139 (38.8%) | 96 / 212 (45.3%) |  |
| 3 | 32 / 139 (23.0%) | 38 / 212 (17.9%) |  |
| 4 | 10 / 139 (7.2%) | 7 / 212 (3.3%) |  |
| 5 | 2 / 139 (1.4%) | 5 / 212 (2.4%) |  |
| Race/Ethnicity |  |  | 0.638 |
| White | 73 / 129 (56.6%) | 109 / 207 (52.7%) |  |
| AfricanAmerican | 17 / 129 (13.2%) | 41 / 207 (19.8%) |  |
| Hispanic | 11 / 129 (8.5%) | 17 / 207 (8.2%) |  |
| Asian | 7 / 129 (5.4%) | 11 / 207 (5.3%) |  |
| Other | 21 / 129 (16.3%) | 29 / 207 (14.0%) |  |
| Weight (kg) | 91.84 +/- 22.05 | 81.25 +/- 23.48 | **<0.001** |
| BMI | 28.65 +/- 6.53 | 29.63 +/- 8.90 | 0.285 |
| Comorbidities |  |  |  |
| HTN | 92 / 131 (70.2%) | 125 / 208 (60.1%) | 0.058 |
| HLD | 62 / 131 (47.3%) | 70 / 208 (33.7%) | **0.012** |
| DM | 30 / 131 (22.9%) | 49 / 208 (23.6%) | 0.889 |
| CAD | 22 / 131 (16.8%) | 24 / 208 (11.5%) | 0.169 |
| PAD | 11 / 131 (8.4%) | 4 / 208 (1.9%) | **0.005** |
| CKD | 4 / 130 (3.1%) | 4 / 208 (1.9%) | 0.497 |
| Stroke/TIA | 60 / 131 (45.8%) | 90 / 208 (43.3%) | 0.648 |
| Tobacco | 73 / 131 (55.7%) | 76 / 208 (36.5%) | **<0.001** |
| Comorbidity score | 2.71 +/- 1.63 | 2.13 +/- 1.43 | **<0.001** |
| Medications |  |  |  |
| Plavix | 15 / 131 (11.5%) | 33 / 208 (15.9%) | 0.256 |
| Aspirin | 122 / 131 (93.1%) | 200 / 208 (96.2%) | 0.214 |
| Statin | 62 / 131 (47.3%) | 78 / 208 (37.5%) | 0.074 |
| Anastomosis sites | 1.25 +/- 0.45 | 1.32 +/- 0.48 | 0.191 |
| Bypass type |  |  | **0.005** |
| STA-MCA | 104 / 141 (73.8%) | 152 / 215 (70.7%) |  |
| Graft-MCA | 6 / 141 (4.3%) | 32 / 215 (14.9%) |  |
| Other-anterior | 10 / 141 (7.1%) | 14 / 215 (6.5%) |  |
| Other-posterior | 21 / 141 (14.9%) | 17 / 215 (7.9%) |  |
| **Intraoperative Measurements** |  |  |  |
| Cut flow | 59.24 +/- 28.81 | 51.09 +/- 28.78 | **0.028** |
| Bypass flow | 51.10 +/- 36.28 | 52.10 +/- 37.14 | 0.311 |
| Cut flow index | 0.89 +/- 0.49 | 0.85 +/- 0.51 | 0.574 |
| **QMRA Measurements** |  |  |  |
| NOVA first flow | 76.25 +/- 37.18 | 65.57 +/- 40.11 | 0.107 |
| NOVA last flow | 51.71 +/- 39.36 | 43.36 +/- 29.51 | 0.268 |
| Pulsatility index | 0.32 +/- 0.23 | 0.31 +/- 0.24 | 0.911 |
| Resistivity index | 0.25 +/- 0.14 | 0.25 +/- 0.16 | 0.998 |
| Wall shear stress | 1.38 +/- 0.72 | 1.41 +/- 0.95 | 0.812 |
| Bypass diameter | 2.89 +/- 0.63 | 2.86 +/- 0.60 | 0.813 |

**Table 3.** Characteristics of patients with STA-MCA bypasses only, excluding graft usage, comparing male and female. mRS = modified Rankin scale; BMI = body mass index; HTN = hypertension; DM = diabetes mellitus; CAD = coronary artery disease; PAD = peripheral arterial disease; CKD = chronic kidney disease; TIA = transient ischemic attack; STA = superficial temporal artery; MCA = middle cerebral artery; NOVA = non-invasive optimal vessel analysis.

|  | Male (n=104) | Female (n=152) | p-value |
| --- | --- | --- | --- |
| Age | 47.67 +/- 18.03 | 45.22 +/- 16.58 | 0.266 |
| Length of follow-up (year) | 1.97 +/- 2.60 | 2.23 +/- 2.77 | 0.455 |
| Bypass patent (at last follow-up) | 84 / 101 (82.2%) | 107 / 152 (71.1%) | **0.044** |
| Diagnosis |  |  | **0.005** |
| Aneurysm | 17 / 104 (16.3%) | 25 / 152 (16.4%) |  |
| Moyamoya | 37 / 104 (35.6%) | 85 / 152 (55.9%) |  |
| Atherosclerotic | 48 / 104 (46.2%) | 40 / 152 (26.3%) |  |
| Other | 0 / 104 (0.0%) | 2 / 152 (1.4%) |  |
| mRS at surgery |  |  | **0.020** |
| 0 | 4 / 103 (3.9%) | 6 / 149 (4.0%) |  |
| 1 | 28 / 103 (27.2%) | 40 / 149 (26.8%) |  |
| 2 | 44 / 103 (42.7%) | 69 / 149 (46.3%) |  |
| 3 | 19 / 103 (18.4%) | 32 / 149 (21.5%) |  |
| 4 | 8 / 103 (7.8%) | 0 / 149 (0.0%) |  |
| 5 | 0 / 103 (0.0%) | 2 / 149 (1.3%) |  |
| Race/Ethnicity |  |  | 0.938 |
| White | 46 / 92 (50.0%) | 74 / 143 (51.7%) |  |
| AfricanAmerican | 15 / 92 (16.3%) | 27 / 143 (18.9%) |  |
| Hispanic | 9 / 92 (9.8%) | 12 / 143 (8.4%) |  |
| Asian | 6 / 92 (6.5%) | 10 / 143 (7.0%) |  |
| Other | 16 / 92 (17.4%) | 20 / 143 (14.0%) |  |
| Weight (kg) | 93.05 +/- 22.31 | 81.50 +/- 23.31 | **<0.001** |
| BMI | 29.32 +/- 6.67 | 30.55 +/- 8.90 | 0.265 |
| Comorbidities |  |  |  |
| HTN | 63 / 94 (67.0%) | 79 / 144 (54.9%) | 0.062 |
| HLD | 42 / 94 (44.7%) | 51 / 144 (35.4%) | 0.152 |
| DM | 24 / 94 (25.5%) | 42 / 144 (29.2%) | 0.540 |
| CAD | 14 / 94 (14.9%) | 16 / 144 (11.1%) | 0.390 |
| PAD | 9 / 94 (9.6%) | 4 / 144 (2.8%) | 0.024 |
| CKD | 2 / 93 (2.2%) | 4 / 144 (2.8%) | 0.764 |
| Stroke/TIA | 50 / 94 (53.2%) | 76 / 144 (52.8%) | 0.950 |
| Tobacco | 58 / 94 (61.7%) | 44 / 144 (30.6%) | **<0.001** |
| Comorbidity score | 2.81 +/- 1.71 | 2.19 +/- 1.54 | **0.004** |
| Medications |  |  |  |
| Plavix | 9 (9.6%) | 27 (18.8%) | 0.053 |
| Aspirin | 91 (96.8%) | 140 (97.2%) | 0.853 |
| Statin | 45 (47.9%) | 62 (43.1%) | 0.465 |
| Anastomosis sites | 1.16 +/- 0.39 | 1.18 +/- 0.39 | 0.562 |
| **Intraoperative Measurements** |  |  |  |
| Cut flow | 59.68 +/- 29.25 | 50.87 +/- 28.35 | **0.023** |
| Bypass flow | 47.22 +/- 27.29 | 41.20 +/- 24.31 | 0.077 |
| Cut flow index | 0.87 +/- 0.48 | 0.85 +/- 0.51 | 0.785 |
| **QMRA Measurements** |  |  |  |
| NOVA first flow | 72.83 +/- 33.54 | 60.79 +/- 33.93 | **0.043** |
| NOVA last flow | 47.73 +/- 36.20 | 39.95 +/- 26.67 | 0.281 |
| Pulsatility index | 0.33 +/- 0.23 | 0.32 +/- 0.25 | 0.888 |
| Resistivity index | 0.26 +/- 0.14 | 0.26 +/- 0.16 | 0.953 |
| Wall shear stress | 1.41 +/- 0.72 | 1.44 +/- 0.97 | 0.839 |
| Bypass diameter | 2.83 +/- 0.52 | 2.79 +/- 0.50 | 0.623 |

### Sex-Related Differences in Cardiovascular Risk Factors

Medical comorbidities were seen at higher rates in men, with comorbidity score 2.7 vs. 2.1 (p<0.001). Men were noted to have significantly higher rates of hyperlipidemia (p=0.012), peripheral arterial disease (p=0.005), and tobacco use (p<0.001). Although non-significant, men also tended to have higher rates of hypertension, coronary artery disease, and chronic kidney disease. No significant differences were observed, however, in baseline antiplatelet or statin therapy usage between men and women, although women tended to use antiplatelet therapy more frequently and men had higher rates of documented statin use.

### Sex-Related Differences in Hemodynamic Parameters

For all bypasses, cut flow was higher in men 59.2 vs. 51.1 (p=0.028), but no significant differences were seen in intraoperative bypass flow, cut flow index (CFI), follow-up QMRA, or bypass diameter (**Table 2**). After removing cases using interposition grafts, similar differences were redemonstrated, including lower cut flow in women (59.7 vs. 50.9 mL/min, p=0.023) and additionally reduced initial QMRA flow measurements in women during the repeat analysis (72.8 vs. 60.8 mL/min, p=0.043) (**Table 3**). Wall shear stress, pulsatility index, and resistivity index also did not significantly differ between men and women in both analyses.

### Hemodynamic Parameters per Bypass Patency and Patient Sex

Using a 2-way ANOVA, while there was a significant difference in CFI among bypasses that were patent vs. occluded, there were no differences between men and women, and there was no interaction between patient sex, CFI, and bypass patency (**Figure 1**). Bypass diameter, when measured on QMRA, also showed differences between patent and occluded bypasses, but in contrast, there was a sex-specific interaction, particularly for STA-MCA bypasses. Interestingly, bypass diameter in women was similar in both patent and occluded bypasses, but bypass diameter in men was larger in patent bypasses. However, no differences were observed between men and women and bypass patency among the following hemodynamic QMRA-based parameters: wall shear stress, pulsatility index, and resistivity index.

**Figure 1.**
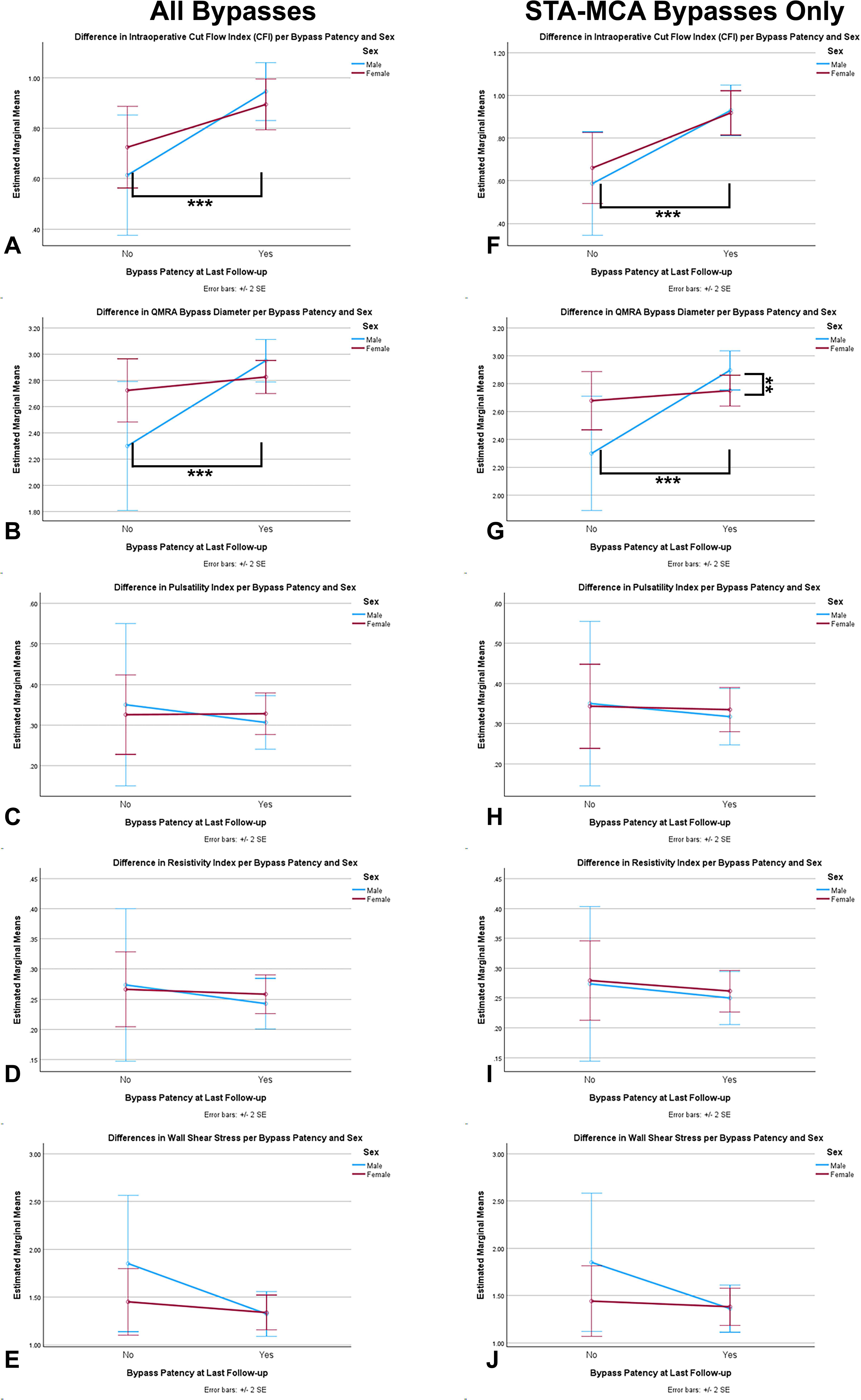
2-way ANOVA comparison of patients with occluded vs. patent bypasses at last follow-up, per patient sex in all bypasses (A-E) and STA-MCA bypasses only (F-J). ** indicates p≤0.05, *** indicates p≤0.005. A) Cut flow index (CFI) significantly different per bypass patency (p=0.002) but no sex-related interaction (p=0.321). B) Bypass diameter measured on QMRA significantly different per bypass patency (p=0.011) but no sex-related interaction (p=0.062). C) No significant difference in pulsatility index per patency (p=0.729) nor sex (p=0.981). D) No significant difference in resistivity index per patency (p=0.604) nor sex (p=0.913). E) No significant difference in wall shear stress per patency (p=0.363) and no interaction with sex (p=0.329). F) CFI significantly different per bypass patency (p<0.001) but no sex-related interaction (p=0.611). G) Bypass diameter measured on QMRA significantly different per bypass patency (p=0.007) and with a significant sex-related interaction (p=0.035). H) No significant difference in pulsatility index per patency (p=0.739) nor sex (p=0.932). I) No significant difference in resistivity index per patency (p=0.597) nor sex (p=0.825). J) No significant difference in wall shear stress per patency (p=0.214) and no interaction with sex (p=0.330).

### Propensity Score-Matched Analysis of Bypass Patency

After propensity score matching, no significant differences were identified between men and women, 228 patients remained in this analysis. Propensity score matching of comorbidities found females have 3.17 higher chance of bypass occlusion (p<0.001, 95% CI 1.60-6.30). When adjusting for CFI, however, the significance of this effect diminishes, but patient sex remained an independent predictor: higher CFI predicts bypass patency (OR 3.16, p=0.028, 95% CI: 1.13-8.81), and females have 2.71 higher odds of bypass occlusion than men (OR 2.71, p=0.017, 95% CI: 1.19-6.18) (**Table 4**).

**Table 4.**
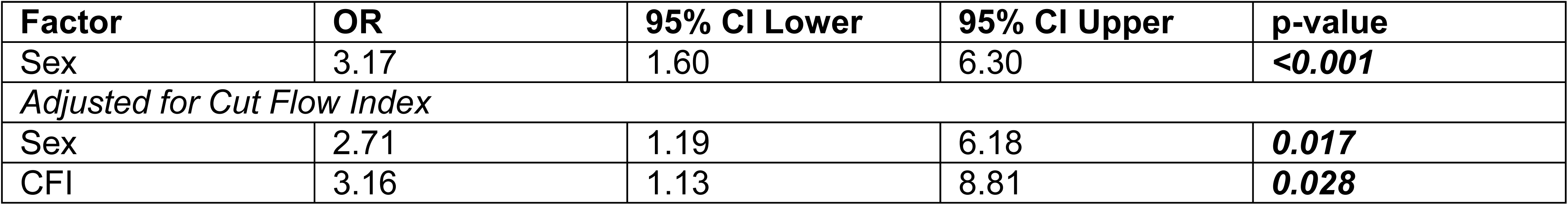
Propensity score-matched analysis of bypass patency. OR = odds ratio; CI = confidence interval; CFI = cut flow index.

## DISCUSSION

This study examined cardiovascular risk factors to assess their impact on cerebrovascular bypass patency and ultimately uncovered a sex-related difference, which has not been previously reported in the neurosurgical literature. Intuitively, one might suspect that cardiovascular risk factors, such as hypertension, hyperlipidemia, diabetes mellitus, and smoking history, might lead to higher rates of bypass occlusion; however, we did not find this to be true. In fact, our initial analysis, seen in **Table 1**, showed that patient sex was the only patient-specific factor that was significantly different between patent and occluded bypasses. Thus, subsequent analyses focused on the examination of biological sex and yielded several interesting findings. First, men generally carried higher rates of comorbidities and cardiovascular risk factors; nonetheless, women showed higher rates of occlusion. This finding was confirmed with a propensity score-matched analysis of cardiovascular risk factors and medical comorbidities, which demonstrated a 3.21 higher odds of bypass occlusion in females.

Next, men and women showed similar flow-based measurements and hemodynamic parameters. Zhao et al. demonstrated higher wall shear stress in women compared to men when examining the large arteries of the head and neck with QMRA.^25^ Alwatban et al. found that the rate of MCA velocity decline and flow pulsatility index rise was significantly greater in females compared with in males.^26^ One could therefore posit that hemodynamic differences in the receiving vascular bed could explain differences in bypass patency, as it relates to sex. However, the findings of our study did not support this theory. Rather, there were no observed differences in wall shear stress, pulsatility index, and resistivity index in patent vs. occluded bypasses, in male vs. female, or in consideration of both factors in the 2-way ANOVA.

Closer inspection with 2-way ANOVA (**Figure 1**) was conducted to assess possible sex-related differences in bypass patency across other hemodynamic parameters. Interestingly, bypass diameter did significantly differ for bypass patency with a sex-related interaction: in women, bypass diameter was similar in both patent and occluded bypasses, whereas in men, bypass diameter was larger in patent bypasses and smaller in occluded bypasses. In addition, cut flow index, which has been described in the literature as a consistent predictor of long-term bypass patency,^15–16,27^ was unsurprisingly generally found to be a significant predictor of bypass patency in this analysis also; however, no difference in CFI was observed between male and female patients. Thus, low CFI predicts occlusion, and high CFI predicts patency at equal rates for men and women. Notably, in the propensity score-matched analysis, when including cardiovascular risk factors and also adjusting for CFI, the impact of sex on bypass patency diminished but still remained independent risk factor outside of CFI.

In total, these findings suggest a possible sex-related contribution to bypass success. Major studies of intracranial bypass procedures exhibit a surprising paucity of information pertaining to patient sex.^28–31^ The most recent publication from the Carotid and Middle Cerebral Artery Occlusion Surgery Study (CMOSS) at least recognizes the need to recruit more women in their study.^30^ Yoon et al., 2019, did report that bypass occlusion did not correlate with patient sex; however, of the 12 bypasses reported as occluded in their study, 9 were women, emphasizing the role of sample size in detecting this difference.^31^

Because this linkage to patient sex and cerebral bypass outcomes has not been previously reported, one can draw from the literature on other vascular diseases. The sex difference in outcomes after coronary artery bypass graft (CABG) has been well established, and when compared to men, women have been shown to have worse operative mortality and morbidity.^32–33^ It is unclear, however, if bypass graft failure rates are higher in women than men, as the literature remains somewhat controversial with some studies finding elevated rates of bypass failure in women^34^ but others not replicating this result.^35^ Worsened outcomes have been seen following carotid endarterectomy in women as well.^36–40^ In the systematic review by den Hartog et al., 2010, the authors identify higher embolic potential in women and differences in plaque morphology as contributors to this difference.^36^

### Limitations

The generalizability of the findings may be limited due to the nature of the study as an observational, retrospective single-center study, and the patients represented in this study primarily reflect an urban, inner-city demographic. The reported rates of bypass occlusion in this study are relatively higher compared to the literature which likely reflects three major differences in this study compared to others: first, a relatively long period of follow-up; second, our database includes all patients where direct bypass was attempted (including failed direct bypasses intraoperatively); and third, different imaging modalities used to determine ongoing patency of the direct bypass (e.g., catheter angiography and QMRA). In addition, the interpretation of CFI in prediction of bypass patency has been studied primarily in steno-occlusive disease^15–16,27^; thus, its inclusion to understand bypass patency in all indications in this study, including both flow augmentation and flow replacement bypasses, would need additional validation. Ultimately, further, larger, prospective, and multi-center studies would be required to confirm these findings.

## CONCLUSION

The findings of this study potentially implicate biological sex in long-term bypass patency, across diagnoses. Women were significantly less likely to have patent bypasses at last follow-up, despite having less medical comorbidities than men and despite having similar intraoperative cut flow index and QMRA flow-related measurements, including bypass flow, pulsatility index, resistivity index, and wall shear stress. In fact, in our analysis, patient sex remained an independent predictor of bypass patency, even after adjusting for CFI and propensity matching. Importantly, no studies in the neurosurgical literature presently describe sex-related differences in cerebral bypass, although sample size may be a limiting factor in prior publications. Further study would be warranted to better elucidate the influence of sex on long-term bypass patency.

## Data Availability

Data may be made available upon appropriate request.

## ABBREVIATIONS

BMI: Body mass index
CABG: Coronary artery bypass graft
CAD: Coronary artery disease
CKD: Chronic kidney disease
CI: Confidence interval
CFI: Cut flow index
DM: Diabetes mellitus
HTN: Hypertension
MCA: Middle cerebral artery
MMD: MoyaMoya Disease
mRS: Modified Rankin scale
NOVA: Non-invasive Optimal Vessel Analysis
PAD: Peripheral arterial disease
PI: Pulsatility index
RI: Resistivity index
STA: Superficial temporal artery
TIA: Transient ischemic attack
QMRA: Quantitative magnetic resonance angiography
WSS: Wall shear stress

---

**Supplemental Table 1.**
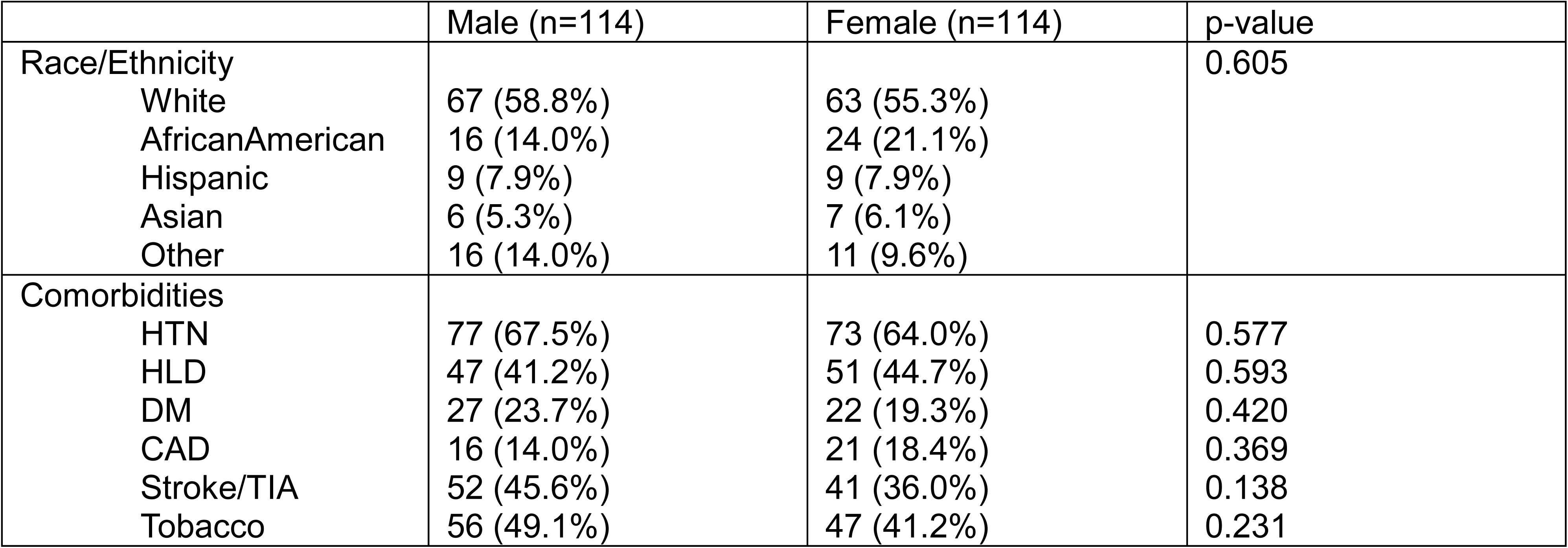
Propensity score-matched variables Characteristics of patients with all bypass types, comparing male and female. HTN = hypertension; DM = diabetes mellitus; CAD = coronary artery disease; TIA = transient ischemic attack.

## REFERENCES

1. Thomas Q, Crespy V, Duloquin G, et al. Stroke in women: When gender matters. Revue Neurologique. 2021/10/01/ 2021;177(8):881–889.

2. Dahl S, Hjalmarsson C, Andersson B. Sex differences in risk factors, treatment, and prognosis in acute stroke. Womens Health (Lond). Jan-Dec 2020;16:1745506520952039.

3. Kumar A, McCullough L. Cerebrovascular disease in women. Ther Adv Neurol Disord. 2021;14:1756286420985237.

4. Vlak MH, Algra A, Brandenburg R, Rinkel GJ. Prevalence of unruptured intracranial aneurysms, with emphasis on sex, age, comorbidity, country, and time period: a systematic review and meta-analysis. Lancet Neurol. Jul 2011;10(7):626–36.

5. Fréneau M, Baron-Menguy C, Vion AC, Loirand G. Why Are Women Predisposed to Intracranial Aneurysm? Front Cardiovasc Med. 2022;9:815668.

6. Peters SAE, Carcel C, Millett ERC, Woodward M. Sex differences in the association between major risk factors and the risk of stroke in the UK Biobank cohort study. Neurology. 2020;95(20):e2715–e2726.

7. Song JW, Xiao J, Cen SY, et al. Sex Differences in Intracranial Atherosclerosis in Patients With Hypertension With Acute Ischemic Stroke. J Am Heart Assoc. May 17 2022;11(10):e025579.

8. Hou L, Li S, Zhu S, et al. Lifetime Cumulative Effect of Reproductive Factors on Stroke and Its Subtypes in Postmenopausal Chinese Women: A Prospective Cohort Study. Neurology. Apr 11 2023;100(15):e1574–e1586.

9. Peters SAE, van der Schouw YT, Woodward M, Huxley RR. Sex Differences in Smoking-related Risk of Vascular Disease and All-cause Mortality. Current Cardiovascular Risk Reports. 2013/12/01 2013;7(6):473–479.

10. Gillis EE, Sullivan JC. Sex Differences in Hypertension. Hypertension. 2016/12/01 2016;68(6):1322–1327.

11. Wang X, Magkos F, Mittendorfer B. Sex Differences in Lipid and Lipoprotein Metabolism: It’s Not Just about Sex Hormones. The Journal of Clinical Endocrinology & Metabolism. 2011;96(4):885–893.

12. Amin-Hanjani S, Alaraj A, Charbel FT. Flow replacement bypass for aneurysms: decision-making using intraoperative blood flow measurements. Acta Neurochir (Wien). 2010;152(6):1021–1032; discussion 1032.

13. Charbel FT, Hoffman WE, Misra M, Ostergren L. Ultrasonic perivascular flow probe: technique and application in neurosurgery. Neurol Res. 1998 Jul;20(5):439–42.

14. Charbel FT, Ed. Flow assisted aneurysm surgery-cerebral aneurysm handbook. New York: Transonic Systems. 2004.

15. Amin-Hanjani S, Du X, Mlinarevich N, et al. The cut flow index: an intraoperative predictor of the success of extracranial-intracranial bypass for occlusive cerebrovascular disease. Neurosurgery. 2005 Jan;56(1 Suppl):75–85; discussion 75-85.

16. Stapleton CJ, Atwal GS, Hussein AE, et al. The cut flow index revisited: utility of intraoperative blood flow measurements in extracranial-intracranial bypass surgery for ischemic cerebrovascular disease. J Neurosurg. 2019 Sep 6:1–5.

17. Alaraj A, Amin-Hanjani S, Shakur SF, Aletich VA, Ivanov A, Carlson AP, Oh G, Charbel FT. Quantitative assessment of changes in cerebral arteriovenous malformation hemodynamics after embolization. Stroke. 2015 Apr;46(4):942–7.

18. Brunozzi D, Hussein AE, Shakur SF, Linninger A, Hsu CY, Charbel FT, Alaraj A. Contrast Time-Density Time on Digital Subtraction Angiography Correlates With Cerebral Arteriovenous Malformation Flow Measured by Quantitative Magnetic Resonance Angiography, Angioarchitecture, and Hemorrhage. Neurosurgery. 2018 Aug 1;83(2):210–216.

19. Amin-Hanjani S, Alaraj A, Calderon-Arnulphi M, Aletich VA, Thulborn KR, Charbel FT. Detection of intracranial in-stent restenosis using quantitative magnetic resonance angiography. Stroke. 2010 Nov;41(11):2534–8.

20. Calderon-Arnulphi M, Amin-Hanjani S, Alaraj A, Zhao M, Du X, Ruland S, Zhou XJ, Thulborn KR, Charbel FT. In vivo evaluation of quantitative MR angiography in a canine carotid artery stenosis model. AJNR Am J Neuroradiol. 2011 Sep;32(8):1552–9.

21. Amin-Hanjani S, Du X, Zhao M, Walsh K, Malisch TW, Charbel FT. Use of quantitative magnetic resonance angiography to stratify stroke risk in symptomatic vertebrobasilar disease. Stroke. 2005 Jun;36(6):1140–5.

22. Shakur SF, Hrbac T, Alaraj A, Du X, Aletich VA, Charbel FT, Amin-Hanjani S. Effects of extracranial carotid stenosis on intracranial blood flow. Stroke. 2014 Nov;45(11):3427–9.

23. Guppy KH, Charbel FT, Corsten LA, Zhao M, Debrun G. Hemodynamic evaluation of basilar and vertebral artery angioplasty. Neurosurgery. 2002 Aug;51(2):327–33; discussion 333-4. PMID: 12182770.

24. Zhao M, Charbel FT, Alperin N, Loth F, Clark ME. Improved phase-contrast flow quantification by three-dimensional vessel localization. Magn Reson Imaging. 2000 Jul;18(6):697–706.

25. Zhao X, Zhao M, Amin-Hanjani S, Du X, Ruland S, Charbel FT. Wall shear stress in major cerebral arteries as a function of age and gender--a study of 301 healthy volunteers. J Neuroimaging. 2015 May-Jun;25(3):403-7.

26. Alwatban MR, Aaron SE, Kaufman CS, Barnes JN, Brassard P, Ward JL, Miller KB, Howery AJ, Labrecque L, Billinger SA. Effects of age and sex on middle cerebral artery blood velocity and flow pulsatility index across the adult lifespan. J Appl Physiol (1985). 2021 Jun 1;130(6):1675–1683.

27. McGuire, LS, Abou Mrad T, Atwal G, Amin-Hanjani S, Charbel FT. Does Disease Etiology Matter in Long-term Patency in Extracranial-Intracranial Bypass? J Neurosurg. 2024 Sept. E-pub ahead of print.

28. EC/IC Bypass Study Group. Failure of extracranial-intracranial arterial bypass to reduce the risk of ischemic stroke. Results of an international randomized trial. N Engl J Med. 1985 Nov 7;313(19):1191–200. doi: 10.1056/NEJM198511073131904. PMID: 2865674.

29. Powers WJ, Clarke WR, Grubb RL Jr, Videen TO, Adams HP Jr, Derdeyn CP; COSS Investigators. Extracranial-intracranial bypass surgery for stroke prevention in hemodynamic cerebral ischemia: the Carotid Occlusion Surgery Study randomized trial. JAMA. 2011 Nov 9;306(18):1983–92. doi: 10.1001/jama.2011.1610. Erratum in: JAMA. 2011 Dec 28;306(24):2672. Obviagele, Bruce. PMID: 22068990; PMCID: PMC3601825.

30. Ma Y, Wang T, Wang H, Amin-Hanjani S, Tong X, Wang J, Tong Z, Kuai D, Cai Y, Ren J, Wang D, Duan L, Maimaitili A, Hang C, Yu J, Bai X, Powers WJ, Derdeyn CP, Wu Y, Ling F, Gu Y, Jiao L; CMOSS Investigators. Extracranial-Intracranial Bypass and Risk of Stroke and Death in Patients With Symptomatic Artery Occlusion: The CMOSS Randomized Clinical Trial. JAMA. 2023 Aug 22;330(8):704–714. doi: 10.1001/jama.2023.13390. PMID: 37606672; PMCID: PMC10445185.

31. Yoon S, Burkhardt JK, Lawton MT. Long-term patency in cerebral revascularization surgery: an analysis of a consecutive series of 430 bypasses. J Neurosurg. 2019 Jul 1;131(1):80–87. doi: 10.3171/2018.3.JNS172158. Epub 2018 Aug 24. PMID: 30141754.

32. Harik L, Perezgrovas-Olaria R, Jr Soletti G, Dimagli A, Alzghari T, An KR, Cancelli G, Gaudino M. Sex differences in coronary artery bypass graft surgery outcomes: a narrative review. J Thorac Dis. 2023 Sep 28;15(9):5041–5054. doi: 10.21037/jtd-23-294. Epub 2023 Aug 16. PMID: 37868858; PMCID: PMC10586965.

33. Alam M, Bandeali SJ, Kayani WT, Ahmad W, Shahzad SA, Jneid H, Birnbaum Y, Kleiman NS, Coselli JS, Ballantyne CM, Lakkis N, Virani SS. Comparison by meta-analysis of mortality after isolated coronary artery bypass grafting in women versus men. Am J Cardiol. 2013 Aug 1;112(3):309–17. doi: 10.1016/j.amjcard.2013.03.034. Epub 2013 May 1. PMID: 23642381.

34. Singh B, Singh G, Tripathy A, Larobina M, Goldblatt J, Tatoulis J. Comparing the patency of the left internal mammary in single, sequential, and Y grafts. J Thorac Cardiovasc Surg. 2024 Jan;167(1):176–182.

35. Lehtinen ML, Harik L, Soletti G, Rahouma M, Dimagli A, Perezgrovas-Olaria R, Audisio K, Demetres M, Gaudino M. Sex differences in saphenous vein graft patency: A systematic review and meta-analysis. J Card Surg. 2022 Dec;37(12):4573–4578.

36. den Hartog AG, Algra A, Moll FL, de Borst GJ. Mechanisms of gender-related outcome differences after carotid endarterectomy. J Vasc Surg. 2010 Oct;52(4):1062–71, 1071.e1-6.

37. Rothwell PM. ACST: which subgroups will benefit most from carotid endarterectomy? Lancet 2004;364:1122–3; author reply 1125-6.

38. Rothwell PM, Eliasziw M, Gutnikov SA, Warlow CP, Barnett HJ; Carotid Endarterectomy Trialists Collaboration. Endarterectomy for symptomatic carotid stenosis in relation to clinical subgroups and timing of surgery. Lancet 2004;363:915–24.

39. Goldstein LB, Samsa GP, Matchar DB, Oddone EZ. Multicenter review of preoperative risk factors for endarterectomy for asymptomatic carotid artery stenosis. Stroke 1998;29:750–3.

40. Sarac TP, Hertzer NR, Mascha EJ, O’Hara PJ, Krajewski LP, Clair DG, et al. Gender as a primary predictor of outcome after carotid endarterectomy. J Vasc Surg 2002;35:748–53.

